# Prevalence and its Associated Factors of Chronic Kidney Diseases among Adult Diabetic Patients in Harari Region, Eastern Ethiopia

**DOI:** 10.1101/2025.08.12.25333507

**Authors:** Temesgen Teshome, Chala Mohammed, Aboma Motuma

**Author notes:** Corresponding author (AM).

## Abstract

Chronic Kidney Disease (CKD) is a major complication of diabetes with a high burden in low-resource settings including Ethiopia due to lack of dialysis, early screening and failure to identify the risk factors for the disease. In Ethiopia, there is a shortage of comprehensive evidence on CKD among diabetic patients. Therefore, this study focused on the gap to addresses CKD in diabetic patients and associated factors in Harari region, eastern Ethiopia. Hospital-based cross-sectional retrospective study was conducted among 315 diabetic patients. Five years data (from January 1, 2019, to December 31, 2024) was collected. Data was collected using Kobo software through chart reviews and checklists in February, 2025, and analyze by SPSS version 25. Descriptive statistics were used to describe frequency, percentage, and tabulation. Bivariable and multivariable logistic regression analyses were conducted to identify associated factors with the outcome variable. Adjusted odds ratio (AOR) with 95% confidence intervals were reported to quantify the strength of associations, and statistical significance was declared at a p-value < 0.05.The study revealed that the prevalence of CKD was (33.2%, 95% CI: 28.5-38.3) among diabetic patients. Alcohol consumption (AOR, 3.89; 95% CI: 1.88-8.03), duration of diabetic (AOR, 3.13; 95% CI: 1.62, 3.77), LDL (AOR, 2.13; 95% CI: 1.21-2.32), prior renal disease (AOR, 4.51; 95% CI: 4.36-5.45), and history of cardiovascular disease (AOR, 2.28; 95% CI: 1.17-4.43) were significantly associated with CKD. However, oral antidiabetic medications has protective effect of CKD (AOR, 0.025; 95% CI: 0.007-0.094). In the study, more than one-third of diabetic patients has developed CKD. Alcohol consumption, LDL, duration of diabetic, prior renal disease, history of cardiovascular disease, and oral therapy were found significantly associated with CKD. A tailored interventions like screening, and awareness creation should be given to diabetic patients to reduce the burden of CKD among diabetic patients.

## Introduction

Chronic kidney disease is defined as structural or functional abnormalities of the kidney that persisted for more than 3 months (1). The international guideline defines CKD as either a decrease in eGFR of <60 ml/min/1.73 m² or presence of markers of kidney damage or both, of at least 3 months duration (2). CKD is a major complication of diabetes mellitus and significantly contributes to morbidity and mortality in the worldwide. The global prevalence of CKD is estimated to be about 11-13% with the majority of cases are contributed from low to middle-income countries (3). Globally it affects around 13.3 million people annually with 85% of the cases are from developing countries (4, 5).The increased rates of diabetes facilitated and escalated chronic kidney disease-related morbidity and mortality(6). Diabetes mellitus is a chronic hyperglycemic state resulting from either insulin deficiency or insulin resistance (7). The incidence of diabetic kidney disease ranged from 20-58/1000 per year(8). Its complications are mostly detected at advanced stages. So early screening of diabetic patients is cost-effective and it helps us to plan early intervention (9).

Diabetes is the leading cause of CKD (10). In 2017 global data show that around 700 million people were having CKD which accounted for 9.1% of the global population and prevalence, incidence and mortality was increased by 29.3%, 43.1% and 41.5% respectively since 1990 (11, 12). In 2019, global burden of disease study show that about 139.6 million people with diabetes developed CKD and about 414.2 thousand of them were died(12).

The prevalence of diabetes is increasing in Africa particularly in Sub-Saharan region with a parallel increase in the prevalence of diabetic kidney disease from 16.3% to 28.3% in 2015 (11). The mortality rate is also increasing due to the lack of dialysis, early screening and failure to identify the risk factors for the disease (13).

There is also lack of early screening for diabetic nephropathy that the patients are diagnosed at advanced stage. Patients at advanced stage of the disease are usually in need of dialysis which isn’t widely available and worsens the situation in the country. As a result of this, identifying the perpetuating risk factors and early intervention is cost effective to reduce the morbidity and mortality rate from chronic kidney disease (14). Most of the previous studies identified that advanced age, long duration of diabetes, comorbid hypertension and poor glycemic status as strong risk factors for diabetic kidney disease (15).

Even if there had been few published reports on the prevalence of CKD and associated factors in diabetic patients in different regions of Ethiopia, they hadn’t assessed the impacts of socio-demographic, lipid profile, and clinical characteristics, substance use and body mass index on the progression to the disease. This study assessed the last five years situation of CKD including the use of both functional (eGFR) and structural (imaging) assessments to diagnose CKD, and enhancing diagnostic accuracy. Additionally, considering a wide range of potential risk factors provided a comprehensive analysis of CKD determinants among diabetic patients and associated risk factors in the study area. It would be used by policymakers in the health sector to develop possible strategies and to design effective programs to identify risk factors and predictors of adverse disease outcomes. The result of this study would have a paramount benefit for the patients, health care workers including clinicians who directly care for the patients and public health personnel, health institutions, and the public hospitals.

## Materials and methods

### Study area, design, and study period

The study was conducted in Harari regional state, Harar city eastern Ethiopia, which is 526 km away from capital city of Ethiopia, Addis Ababa. According to central statistics agency (CSA), the region has an estimated area of 340 km^2^ with 19 urban and 17 rural kebeles (the smallest administrative unit structure). It was populated region, 183,344 peoples with an equal ratio of male to female population. About 99,321 people were living in urban area and the remaining 84,023 were rural inhabitants (16). According to the current regional health bureau, there are two public Hospitals (Hiwot Fana Comprehensive Specialized University Hospital & Jugal Hospital), one Federal Police Hospital, one private Hospital (Harar General Hospital), nine Health Centers, twenty-nine private clinics, twenty-seven health posts and one regional laboratory in the region (17).

Hospital based cross-sectional retrospective study was conducted in Hiwot-Fana Comprehensive Specialized University Hospital (HFCSUH) and Jugal General Hospital (JGH). The study was conducted by reviewing medical records of diabetic patients who visited Hiwot Fana Comprehensive Specialized University Hospital and Jugel General Hospital from January 1, 2019 to December 31, 2024, Harari region, Eastern Ethiopia. The data were collected from February 1 to 30, 2025.

### Inclusion and exclusion criteria

All adult type 2 and type 1 diabetic patients whose age ≥18 years and complete records of information on study variables were included. In addition, patients who visited diabetic clinic in the public hospitals of Harari region from January 1, 2019 to December 31, 2024 were eligible for the study. Patients with urinary tract obstruction, bilateral renal artery stenosis and active urinary sediment (hematuria, casts) was excluded from the study.

### Sample size determination

The sample size was determined using a single population proportion formula, taking the prevalence of CKD in diabetic patients from a study done in Dessie Referral Hospital, South Wollo, Ethiopia (32%) (18), using margin of error (d)= 5%, the sample size was determined as follows

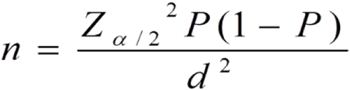

Zα/2=1.96, CI= 95% confidence interval), calculated ample size (n) = 334 and adding 5% for non-response rate, then the final sample size was 351

### Sampling procedure and technique

The registry log book of patients who visited Hiwot fana Comprehensive Specialized Hospital and Jugel General Hospital were used. Then, the sample size for each hospital was allocated based on the proportion of the diabetic patients registered at diabetic clinic. Lastly, the study participants were chosen from each hospital sample frame using the medical record numbers (MRN) of diabetic patients were taken and the charts were randomly selected using simple random sampling technique until the required sample size were fulfilled the requirement numbers.

### Data collection methods

The HMIS registration logbook was used to get the card numbers of diabetic patients who had visited the hospitals during the study period. After getting the card numbers, patient charts were retrieved from the card office, and data were collected with Kobo Collect application. A retrospective chart review was conducted. Structured checklists were prepared by the principal investigator based on literature reviews. Structured checklists were used to extract data from patients’ charts. It was pre-tested, and obtain demographic data, medical, behavioral and measured laboratory factors. Data collection was performed by five medical interns (final year medical students) who were off duty during data collection time. All the data collectors were trained about the objectives of the study and data were extracted from patients’ chart by the data collectors under the supervision of the principal investigator.

### Dependent and independent variables

The outcome variable was CKD, and independent variables ware sociodemographic variables (age, gender, residency, educational status, occupational and marital status), Medical and behavioural factors like smoking, khat chewing, obesity, hypertension, type of diabetes, family history of chronic kidney disease, alcohol consumption, previous history of kidney disease, duration of diabetes, diabetic retinopathy, nonsteroidal anti-inflammatory drug use, type of medication and adherence, comorbid cardiovascular disease, and laboratory tests (serum lipid level, HgA1C and FBS level, degree of proteinuria).

### Operational definitions

Chronic Kidney Disease is decreased kidney function shown by eGFR of less than 60mL/min/1.73 m2, and/or markers of kidney damage of at least 3 months duration regardless of the underlying cause (19).

Diabetes mellitus: If the patient is either previously on anti-diabetic medications, or had diagnosis of any type of diabetes, or fasting blood sugar >=126 mg/dl or RBS >=200 mg/dl or HgA1C of ≥6.5% (20)

Hypertension: is defined whenever newly or previously detected blood pressure ≥140/90mmHg persisting for two different measurements at least one week apart (21).

Dyslipidemia or hyperlipidemia: is considered whenever the serum total cholesterol is ≥200mg/dl, LDL-C≥100mg/dl, triglyceride level≥150mg/dl, HDL-C˂ 40mg/dl for men and ˂50mg/dl for women or previous diagnosis of hyperlipidaemia (22)

Estimated Glomerular filtration rate (eGFR): is calculated based on height, weight, gender, and serum creatinine level for assessment of kidney function (23).

Fasting blood sugar (FBG): Serum glucose level measured after at least 8 h of fasting (24) Overweight is defined whenever the BMI is≥ 25-29.9 kg/m2, while obesity is defined whenever the BMI is ≥30 kg/m^2^ (25).

### Data quality control

To maintain data quality, it was ensured at every step of the study from data collection and coding to entry, cleaning, and analysis. The data collectors and supervisors underwent two days of training, which focused on the study’s relevance, objective, ethical issues, informed consent, and interviewing techniques. The questionnaire was pretested in different sites other than those selected for the study before the actual data collection. The checklists were pretested and its language clarity were revised. The investigator supervised the data collection process on daily based. .

### Methods of data analysis

Data was entered using Kobo Collect software and analyzed using SPSS version 25. Descriptive analysis was used to describe frequency and percentage for categorical variables; and mean and standard deviation for continuous variables were used to describe the characteristics of study subjects. Chi square test with p-value was used when appropriate to test the significance of changes in these variables. Binary logistic regression model was used to determine the relationship between the outcome and predictor variables. Bivariable logistic regression analyses was carried out to assess the association between the dependent and independent variables and to identify candidates for multivariable analysis. Then, multivariable analysis was performed on the variables which have a P-value <0.25 to determine the independent predictors of CKD. Multicollinearity was checked. The backward regression was fitted and model fitness was checked by the Hosmer-Lemeshow test (P=0.0756). Statistical significance was measured by p-value < 0.05 and adjusted odds ratio (AOR) with a 95% confidence interval (95% CI).

### Ethical considerations

Ethical clearance was obtained from the Haramaya University, College of Health and Medical Sciences Institutional Health Research Ethics Review Committee (Ref. No. IHRERC/021/2025) prior to data collection. Informed voluntary written and signed consent was obtained from heads of the hospitals and the letter was submitted to all responsible bodies to ensure smooth and effective data collection. Information gained from medical records was kept as anonymous and confidential.

## Results

### Socio-demographic characteristics of study participants

This study was conducted on 351 adult diabetic participants with their mean age of 60.9 and (SD=12.23) years. The majority of the study participant was female which accounted for 51.42% (n= 181), and rural residents accounted for 63.07% (n= 221). In this study, about 58.89% (n=207) of the participants were unemployed. Regarding the marital status, about 75.28% (n= 264) of the participants were married; 6.25% (n= 22) of them were divorced; 17.61% (n= 62) were widowed; and the remaining 0.85% (n= 3) of the participants were single. In terms of educational status, the one-third of the study participants had attended either primary school (31.25%, n= 110), secondary school (19.03%, n= 67) or college/University (39.49%, n= 139) (Table 1).

**Table 1.** Sociodemographic characteristics of study participants among adult diabetic patients who visited public hospitals of Harari Region, Eastern Ethiopia, 2024 (N=351)

| Variables |  | Frequency (n) | Percentage (%) |
| --- | --- | --- | --- |
| Age of mean (SD) |  | 60.88 (SD=12.23, Min=25, Max=89) |  |
| Gender | Female | 181 | 51.42 |
|  | Male | 170 | 48.58 |
| Residence | Rural | 221 | 63.07 |
|  | Urban | 130 | 36.93 |
| Employment status | Unemployed | 207 | 58.81 |
|  | Employed | 144 | 41.19 |
| Marital status | Married | 264 | 75.28 |
|  | Widowed | 62 | 17.61 |
|  | Divorced | 22 | 6.25 |
|  | Single | 3 | 0.85 |
| Educational status | College/University | 138 | 39.49 |
|  | Primary education | 110 | 31.25 |
|  | Secondary education | 67 | 19.03 |
|  | No Formal education | 36 | 10.23 |

### Behavioral characteristics

Self-reported behavioral lifestyle of the study participants show that, the majority of participants had a history of khat chewing (85.23%, n =300) and smoking (53.13%, n=186), and about 74.43%, n=261) had no alcohol consumption (Table 2).

**Table 2.** Substance use behavior of study participants among adult diabetic patients who visited public hospitals of Harari Region, Eastern Ethiopia, 2024 (N=351)

| Variables | Categories | Frequency (n) | Percentage (%) |
| --- | --- | --- | --- |
| Smoking history | No | 165 | 44.88 |
|  | Yes | 186 | 53.13 |
| History of alcohol consumption | No | 261 | 74.43 |
|  | Yes | 90 | 25.57 |
| Khat Chewing | No | 51 | 14.77 |
|  | Yes | 300 | 85.23 |

### Clinical characteristics

The clinical profile of the participants revealed that several diabetes-related complications and comorbid conditions might contribute to the development of CKD. Notably, a large proportion of the sample exhibited signs of advanced diabetic complications: approximately 65% had retinopathy and 40% had diabetic foot ulcers. Additionally, nearly one-third of the patients reported a history of renal disease (31.53%), while a similar proportion (33.24%) had a family history of renal disease (Table 3).The majority of the participants had type II diabetes (86.36%) with a mean duration of about 15 years. Most of the participants were using oral medications which accounted for 40.06% (n= 140) while about 38.07% (n= 134) of the participants were using combination therapy and the remaining 21.88% (n=77) participants were on injectable therapy. Moreover, about 44.89% (n= 157) of the participants were identified to have a history of cardiovascular disease. Over 80% of the participants were either overweight or obese, (43.47% overweight and 36.65% obese) (Table 3).

**Table 3.** Clinical characteristics of study participants among adult diabetic patients who visited public hospitals of Harari Region, Eastern Ethiopia, 2024 (N=351)

| Variables | Category | Frequency (n) | Percentage (%) |
| --- | --- | --- | --- |
| Retinopathy | No | 22 | 34.94 |
|  | Yes | 229 | 65.06 |
| Diabetic foot ulcer | No | 210 | 59.94 |
|  | Yes | 141 | 40.06 |
| Previous renal disease | No | 241 | 68.47 |
|  | Yes | 110 | 31.53 |
| Family History of renal disease | No | 235 | 66.76 |
|  | Yes | 116 | 33.24 |
| DM Type | Type I | 48 | 13.64 |
|  | Type II | 303 | 86.36 |
| Duration of DM | Mean=15 years (Min=1 Max=35 SD=7.16) |  |  |
| Medication type | Injectable | 77 | 21.88 |
|  | Oral | 140 | 40.06 |
|  | Combination | 134 | 38.07 |
| History of cardiovascular disease | No | 194 | 55.11 |
|  | Yes | 157 | 44.89 |
| BMI | Normal | 70 | 19.89 |
|  | Overweight | 152 | 43.47 |
|  | Obese | 129 | 36.65 |

Legend: DM, Diabetes Mellitus; BMI, Body Mass Index

### Biochemical parameter characteristics

The findings highlighted a considerable burden of renal and metabolic derangements among participants. Notably, the 24-hour or urine dipstick analysis for proteinuria revealed that only a small fraction (3.13%) had no detectable protein, while nearly 75% exhibited moderate to severe proteinuria. This graded increase in proteinuria suggested significant renal involvement, which was consistent with CKD progression (Table 4).Glycemic control was suboptimal in the majority of the participants, with approximately 70% classified as poorly controlled. Moreover, the lipid profile was high among the study participants, for instance, over two-thirds of participants had elevated total serum cholesterol (69.32%) and high levels of serum triglycerides (58.81%) and LDL (56.25%), while low HDL was observed in nearly 77% of the subjects. Such dyslipidemia is known to contribute to endothelial dysfunction and atherosclerosis, potentially accelerating the decline in renal function (Table 4).

**Table 4:** Biochemical profile of study participants among adult diabetic patients who visited public hospitals of Harari Region, Eastern Ethiopia, 2024 (N=351)

| Variables | Category | Frequency (%) | Percentage (%) |
| --- | --- | --- | --- |
| Proteinuria (24 hr urine or dipstick) | No | 11 | 3.13 |
|  | 1 | 73 | 20.74 |
|  | 2 | 156 | 44.32 |
|  | 3 | 105 | 29.83 |
|  | 4 | 6 | 1.99 |
| Glycemic control (FBS)/HgA1C | Poorly controlled | 244 | 69.60 |
|  | Well controlled | 107 | 30.40 |
| Total serum cholesterol | High | 244 | 69.32 |
|  | Normal | 107 | 30.68 |
| Serum triglyceride | High | 207 | 58.81 |
|  | Normal | 144 | 41.19 |
| LDL | High | 197 | 56.25 |
|  | Normal | 154 | 43.75 |
| HDL | Low | 269 | 76.70 |
|  | Normal | 82 | 23.30 |
| Estimated Glomerular Filtration rate (eGFR) | <15 | 35 | 9.94 |
|  | 15-29 | 13 | 3.69 |
|  | 30-44 | 15 | 4.26 |
|  | 45-59 | 53 | 15.34 |
|  | 60-89 | 29 | 8.24 |
|  | >=90 | 206 | 58.52 |
| Imaging evidence of CKD | No | 209 | 59.66 |
|  | Yes | 142 | 40.34 |
| CKD | No | 234 | 66.76 |
|  | Yes | 117 | 33.24 |

**Legends:** CKD, Chronic kidney Diseases; eGFR, Estimated Glomerular Filtration Rate; LDL, Low Density Lipoprotein; HDL, High Density Lipoprotein; FBS, Fast Blood Sugar; HgA1C, HemoglobinA1C

### Prevalence of CKD

The prevalence of the data indicated that more than half of the participants (58.5%) had an eGFR in the normal range (≥90 mL/min/1.73 m²) however a substantial proportion of them exhibited reduced kidney function. Notably, 9.9% had severely reduced function (eGFR <15), 3.7% fell into the 15–29 range, 4.3% into the 30–44 range, 15.3% into the 45–59 range, and 8.2% into the 60–89 range (Fig 1).

**Fig 1.**
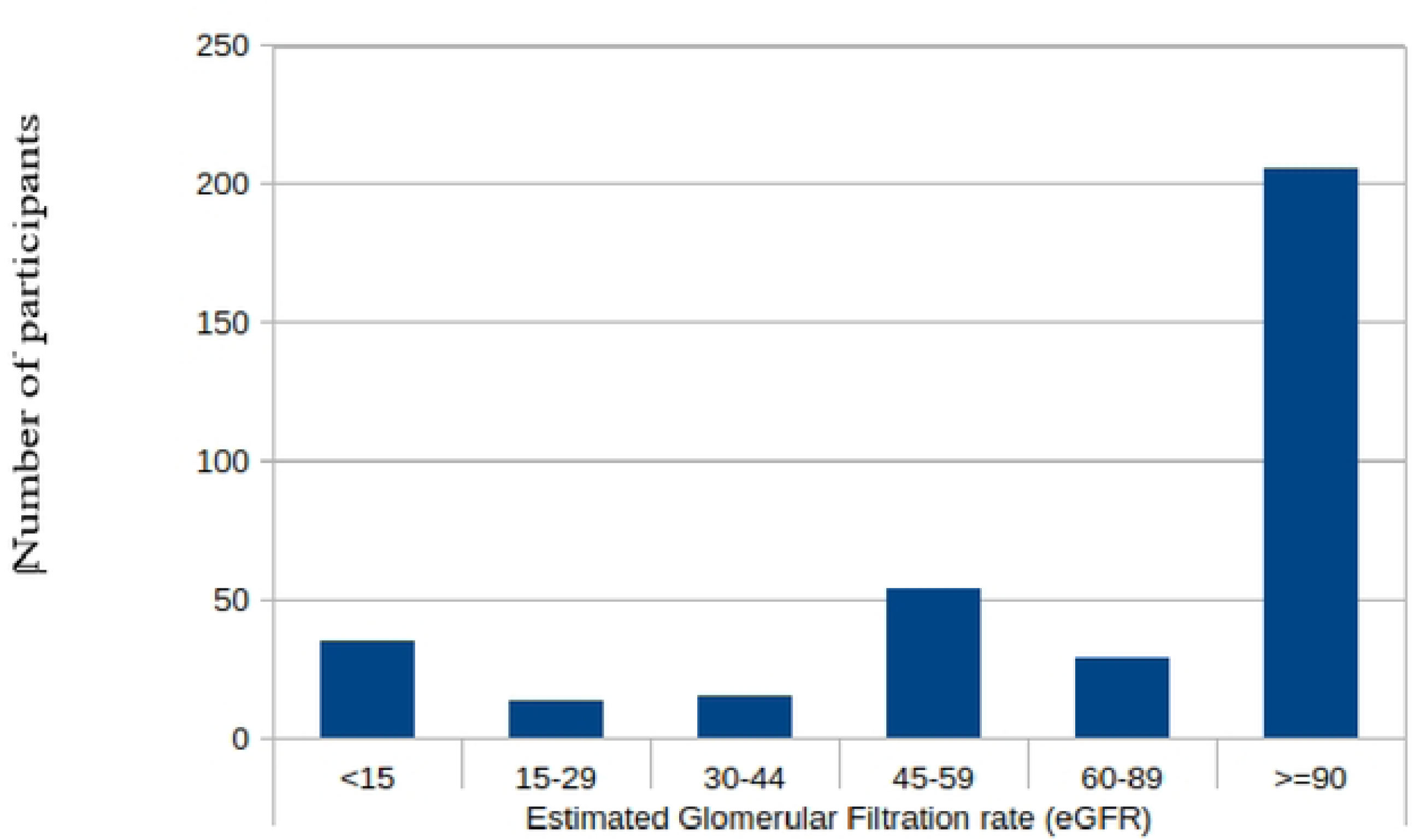
Estimated Glomerular Filtration Rate of Diabetic Patients in Public Hospitals of Harari Region, Eastern Ethiopia, 2019-2024 (N=351)

When considering structural or parenchymal changes, imaging evidence of CKD was observed in 40.3% of the sample. Overall, one-third of the participants (33.2%, 95% CI: 28.5-38.3) met the criteria for CKD. These findings indicated that about one in three diabetic patients were affected by CKD based on functional and imaging assessments.

### Factors associated with CKD

The study revealed that, alcohol consumption (AOR = 3.89; 95% CI: 1.88-8.03), Duration of DM (AOR= 3.13; 95%CI: 1.62, 3.77), previous renal disease (AOR, 4.51; 95% CI: 4.36-5.45), and cardiovascular disease history (AOR = 2.28; 95% CI: 1.17-4.43), LDL (AOR=2.13; 95% CI: 1.21-2.32) were independently associated with CKD. Notably, injectable antidiabetic therapy (insulin) was associated with markedly higher CKD risk compared to oral medications (AOR = 0.03; 95% CI: 0.007-0.094). (Table 5).

**Table 5.**
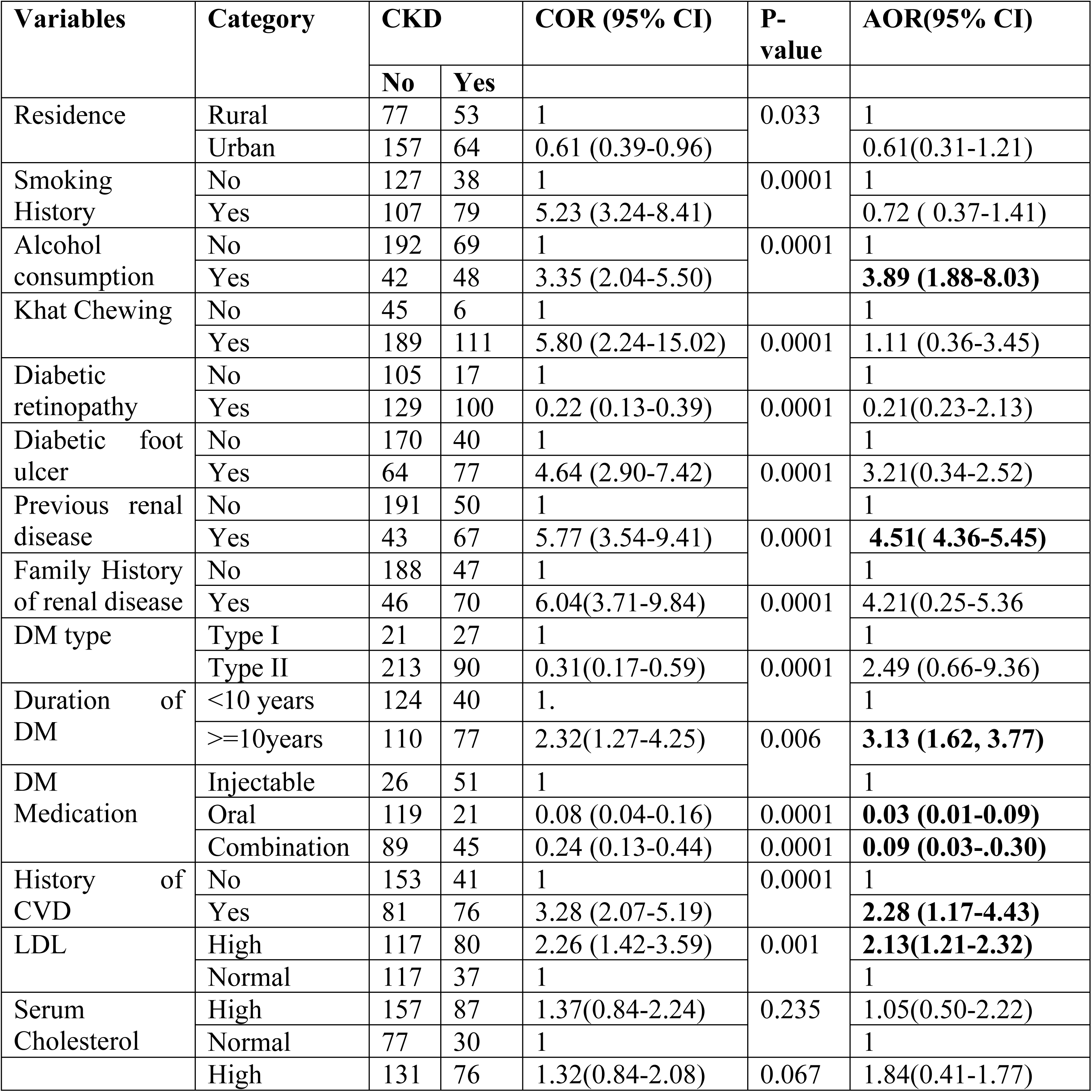
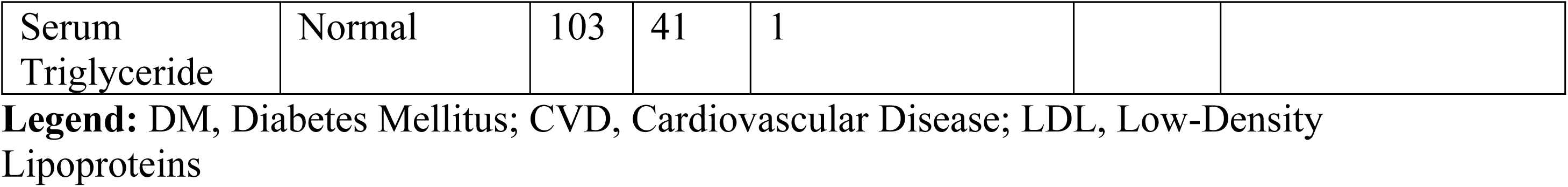
Factors associated with CKD among diabetic patients who visited public hospitals of Harari Region, Eastern Ethiopia, 2024 (N=351)

## Discussion

This study assessed the prevalence and associated factors of CKD among diabetic patients, offering valuable insights into renal health. The findings revealed a substantial burden of CKD, with one-third of participants meeting the diagnostic criteria based on estimated glomerular filtration rate (eGFR) and imaging evidence. Additionally, this study highlighted the influence of lifestyle behaviors, clinical comorbidities, and treatment modalities on CKD risk among diabetic patients.

In this study, the prevalence of CKD was 33.2%, which a substantial disease burden, higher to global estimates CKD approximately 10-16% of the general population worldwide (26), and higher among diabetics patients in princess marina hospital, Gaborone, Botswana 13.5% (27).The American Diabetes Association (ADA) states that 20-40% of diabetic patients develop CKD over time (20). In addition, the high proportion of advanced CKD (9.9% with eGFR <15 mL/min/1.73 m²) exceeds the global average of 5-7% (12), reflecting delayed diagnosis and poor adherence to Kidney Diseases Improving Global Outcome (KDIGO) guidelines for early screening in this population (28). Structural kidney damage 40.3% despite preserved eGFR in many cases highlights the need for combined functional and imaging assessments, as recommended by the International Society of Nephrology (ISN) for high-risk groups (29).

Comparing this study’s findings with existing literature, CKD prevalence among diabetic patients varied across studies and geographic locations. In Ethiopia, Adem et al. (18) reported a prevalence of 31.5% in Dessie Referral Hospital, South Wollo, Ethiopia comparable to our findings. However, a study in Ayder Comprehensive Specialized Hospital, northern Ethiopia reported a lower prevalence of 26.5% (30), 2.7% in pastoralist health facility of Jinka General Hospital, Southern Ethiopia (31), Gondar 21.8% (32), Federal police hospital diabetic clinic in Addis Ababa14.6%(33), and Mekelle 22.1% (34), which may be attributed to differences in study populations, diagnostic criteria, or healthcare access. Moreover, in Ethiopia a recent systematic review and meta-analysis reported that a pooled CKD was 18% among diabetic patients (35).

Furthermore, a study in Harar reported an incidence rate of 2.16 cases per 100 person-years and a 10.32% cumulative incidence over a 10-year follow-up (36). Conversely, studies in high-income countries have reported higher CKD prevalence among diabetics, exceeding 40% in the United States and parts of Europe (37, 38). However, studies in the United Kingdom reported 18.2% (39) and Canada 7.2% (40) reflected better early detection and management, whereas Peru 24.3%(41), and Asian nations 7-34.3% (42) showed variability tied to healthcare infrastructure.

The global prevalence of CKD was estimated of between 11 to 13%(43) , Africa has wide ranges 2-41% (44), with higher rates in East Africa 35.3% for diabetic kidney disease (45), consistent with our findings. Evidence show that in Uganda 33.7% (46) highlight regional disparities, likely due to differences in diabetes care and comorbid burden. These variations may be due to differences in screening practices, genetic predispositions, healthcare infrastructure, and patient adherence to diabetes management.

In Ethiopia, our results (33.2%) fell within the range of prior estimates 9.8-72.7%, though closer to meta-analyses pooling regional data 21.7-35.5% (47), and estimated pooled prevalence of CKD among DM patients in Ethiopia was 18%(35). Discrepancies arise from methodological heterogeneity: Study in Northeast Ethiopia show the prevalence of impaired eGFR was 19%, albuminuria was 30.9%, and 33.9% of the patients had some degree of CKD(48), whereas community-based cohorts study was 15.3% in Dawuro (49) reflect under diagnoses in rural areas. Urban-rural divides are critical, as rural participants (two-thirds of our cohort) face barriers like limited nephrology services (50), whereas urban studies was 32% in Tikur Anbessa Hospital (51) benefit from advanced diagnostics.

In this study, several factors were significantly associated with increased the risk of CKD. For instance, alcohol consumption (AOR = 3.89), Duration of DM (AOR=3.13), prior renal disease (AOR = 4.51), LDL (AOR=2.13), and a history of cardiovascular disease (AOR = 2.28) were independent risk factors. These findings align with studies in Ethiopia (33, 35, 47, 52), Sub-Sahara Africa (53) and globally, which identified cardiovascular comorbidities and pre-existing kidney disease as major contributors to CKD progression. The role of alcohol consumption as a risk factor is also supported by review focused on 21 clinical studies (54, 55), which suggested that excessive alcohol intake exacerbated oxidative stress and renal inflammation, leading to kidney damage. The strong association between cardiovascular disease and CKD was well-documented, as both conditions share common pathophysiological pathways, including endothelial dysfunction and systemic inflammation (56).

Conversely, some factors appeared protective against CKD in our analysis. Urban residence, being overweight and the use of oral antidiabetic medications were associated with lower CKD risk. The protective effect of urban residence, though not statistically significant after adjustment, had been reported in other studies(30) and might be due to better healthcare access and early disease detection. The inverse association between BMI and CKD was an interesting finding, as obesity is generally considered a risk factor for renal dysfunction. However, studies in African populations (47, 57) suggested that being overweight status may provide metabolic reserves that protect against CKD progression. The protective role of oral antidiabetic medications was consistent with studies in Ethiopia (48), and Colombia(58).

Some factors that did not show a significant association with CKD in this study had been reported as risk factors in other studies. Gender, occupation, marital status, and metabolic markers such as glycemic control and lipid profile didn’t independently predict CKD risk. This contradicted with other studies in Ethiopia (31), which found out an association between poor glycemic control and CKD progression. One plausible explanation for this discrepancy was the cross-sectional nature of this study, which limited the ability to establish causality. Additionally, glycemic control and lipid abnormalities might have exerted their effects over a longer period, and variations in study populations, sample sizes, and definitions of metabolic control could contribute to these inconsistencies.

The finding has implication for health care providers and clinical practice. Knowing the burden of CKD among diabetes patients should likely be established prevention, and early screening whenever affordable and feasible. It is used to providing guidance to enhance awareness of CKD among health care professionals and patients and the promotion of healthy lifestyles in preventive programs. Moreover, it provides information about the burden and public health impact of renal failure in the county for possible attention during routine clinical patient care. Furthermore, identifying risk factors may help health care professionals treat DM patients with CKD during their clinical care.

### Strength and limitation of the study

This study is used the five years secondary data to collect as many information as possible to identify CKD and its the determinants of CKD among diabetic patients attending care and follow up at Harar hospitals in the last five years. The study also including the use of both functional (eGFR) and structural (imaging) assessments to diagnose CKD, enhancing diagnostic accuracy. Additionally, considering a wide range of potential risk factors provided a comprehensive analysis of CKD determinants in diabetic patients. However, it has several limitations. Firstly, since the study used cross sectional study design, the causal association would not be inferred. Secondly, the use of secondary data resulted in incompleteness of some variables. Despite this, the study tried to address the main determinants of CKD, clinical morbidity, and lifestyle characteristics of study participants

## Conclusion

This study emphasized a significant burden of CKD among diabetic patients in Ethiopia, with a prevalence of 33.2%. Independent predictors of CKD included alcohol consumption, HDL, duration DM, prior renal disease, and cardiovascular disease, highlighting the independent predictors of CKD among DM patients. However, oral antidiabetic medications were found to be protective. These findings advocate for prioritized public health interventions including reducing alcohol consumption, expanded screening for high-risk groups like those with prior renal disease or CVD, and early adoption of oral antidiabetic regimens.

## Data Availability

All the data will be provided by corresponding author up on the requested

## Acknowledgements

We would like to thank Haramaya University for supporting this research. In addition, we would like to thank data collectors and supervisor for their commitments. We would also like to thank the Harari Regional Health Bureau and their public hospitals for providing the background information, and medical records.

## Author contribution

All authors have equally contributed from the beginning of the proposal conception to the development of this manuscript

## References

1. Navaneethan SD, Zoungas S, Caramori ML, Chan JC, Heerspink HJ, Hurst C, et al. Diabetes management in chronic kidney disease: synopsis of the KDIGO 2022 clinical practice guideline update. Annals of internal medicine. 2023;176(3):381–7.

2. Levey AS, Eckardt K-U, Tsukamoto Y, Levin A, Coresh J, Rossert J, et al. Definition and classification of chronic kidney disease: a position statement from Kidney Disease: Improving Global Outcomes (KDIGO). Kidney international. 2005;67(6):2089–100.

3. Hill NR, Fatoba ST, Oke JL, Hirst JA, O’Callaghan CA, Lasserson DS, et al. Global prevalence of chronic kidney disease–a systematic review and meta-analysis. PloS one. 2016;11(7):e0158765.

4. Lv J-C, Zhang L-X. Prevalence and disease burden of chronic kidney disease. Renal fibrosis: mechanisms and therapies. 2019:3–15.

5. Nazzal Z, Hamdan Z, Masri D, Abu-Kaf O, Hamad M. Prevalence and risk factors of chronic kidney disease among Palestinian type 2 diabetic patients: a cross-sectional study. BMC nephrology. 2020;21(1):484.

6. Jha V, Garcia-Garcia G, Iseki K, Li Z, Naicker S, Plattner B, et al. Chronic kidney disease: global dimension and perspectives. The lancet. 2013;382(9888):260-72.

7. Khunti K, Chudasama YV, Gregg EW, Kamkuemah M, Misra S, Suls J, et al. Diabetes and multiple long-term conditions: a review of our current global health challenge. Diabetes Care. 2023;46(12):2092–101.

8. Gerber C, Cai X, Lee J, Craven T, Scialla J, Souma N, et al. Incidence and progression of chronic kidney disease in black and white individuals with type 2 diabetes. Clinical Journal of the American Society of Nephrology. 2018;13(6):884–92.

9. Kumela Goro K, Desalegn Wolide A, Kerga Dibaba F, Gashe Fufa F, Wakjira Garedow A, Edilu Tufa B, et al. Patient awareness, prevalence, and risk factors of chronic kidney disease among diabetes mellitus and hypertensive patients at Jimma University Medical Center, Ethiopia. BioMed Research International. 2019;2019(1):2383508.

10. Ghaderian SB, Beladi-Mousavi SS. The role of diabetes mellitus and hypertension in chronic kidney disease. Journal of renal injury prevention. 2014;3(4):109.

11. Cheng H-T, Xu X, Lim PS, Hung K-Y. Worldwide epidemiology of diabetes-related end-stage renal disease, 2000–2015. Diabetes Care. 2021;44(1):89–97.

12. Cockwell P, Fisher L-A. The global burden of chronic kidney disease. The lancet. 2020;395(10225):662–4.

13. Deng Y, Li N, Wu Y, Wang M, Yang S, Zheng Y, et al. Global, regional, and national burden of diabetes-related chronic kidney disease from 1990 to 2019. Frontiers in endocrinology. 2021;12:672350.

14. Wagnew F, Eshetie S, Kibret GD, Zegeye A, Dessie G, Mulugeta H, et al. Diabetic nephropathy and hypertension in diabetes patients of sub-Saharan countries: a systematic review and meta-analysis. BMC research notes. 2018;11(1):565.

15. Azagew AW, Abate HK, Mekonnen CK, Mekonnen HS, Tezera ZB, Jember G. Diabetic dyslipidemia and its predictors among people with diabetes in Ethiopia: systematic review and meta-analysis. Systematic Reviews. 2024;13(1):190.

16. CSA. Central Statistical Agency (CSA)[Ethiopia] and ICF. Ethiopia Demographic and Health Survey, Addis Ababa. Central Statistical Agency. 2016.

17. Shama AT, Roba HS, Abaerei AA, Gebremeskel TG, Baraki N. Assessment of quality of routine health information system data and associated factors among departments in public health facilities of Harari region, Ethiopia. BMC Med Inform Decis Mak. 2021;21(1):287.

18. Adem M, Mekonen W, Ausman A, Ahmed M, Yimer A. Prevalence of chronic kidney disease and its associated factors among diabetes mellitus patients in Dessie Referral Hospital, South Wollo, Ethiopia. Sci Rep. 2024;14(1):9229.

19. Webster AC, Nagler EV, Morton RL, Masson P. Chronic Kidney Disease. Lancet. 2017;389(10075):1238–52.

20. ADA. Standards of Care in Diabetes-2023 Abridged for Primary Care Providers. Clinical diabetes : a publication of the American Diabetes Association. 2022;41(1):4–31.

21. Unger T, Borghi C, Charchar F, Khan NA, Poulter NR, Prabhakaran D, et al. 2020 International Society of Hypertension Global Hypertension Practice Guidelines. Hypertension (Dallas, Tex : 1979). 2020;75(6):1334–57.

22. Motuma A, Shiferaw K, Gobena T, Teji Roba K, Berhane Y, Worku A. Dyslipidemia and its predictors among adult workers in eastern Ethiopia: An institution-based cross-sectional study. PLoS One. 2023;18(10):e0291665.

23. Fabian J, George JA, Etheredge HR, van Deventer M, Kalyesubula R, Wade AN, et al. Methods and reporting of kidney function: a systematic review of studies from sub-Saharan Africa. Clinical kidney journal. 2019;12(6):778–87.

24. Yu Q, Mao X, Fu Z, Luo S, Huang Q, Chen Q, et al. Fasting blood glucose as a predictor of progressive infarction in men with acute ischemic stroke. The Journal of international medical research. 2022;50(10):3000605221132416.

25. Donini LM, Busetto L, Bischoff SC, Cederholm T, Ballesteros-Pomar MD, Batsis JA, et al. Definition and Diagnostic Criteria for Sarcopenic Obesity: ESPEN and EASO Consensus Statement. Obes Facts. 2022;15(3):321–35.

26. Deng L, Guo S, Liu Y, Zhou Y, Liu Y, Zheng X, et al. Global, regional, and national burden of chronic kidney disease and its underlying etiologies from 1990 to 2021: a systematic analysis for the Global Burden of Disease Study 2021. BMC Public Health. 2025;25(1):636.

27. Rwegerera GM, Bayani M, Taolo EK, Habte D. The prevalence of chronic kidney disease and associated factors among patients admitted at princess marina hospital, Gaborone, Botswana. Nigerian journal of clinical practice. 2017;20(3):313–9.

28. Stengel B, Muenz D, Tu C, Speyer E, Alencar de Pinho N, Combe C, et al. Adherence to the Kidney Disease: Improving Global Outcomes CKD Guideline in Nephrology Practice Across Countries. Kidney Int Rep. 2021;6(2):437–48.

29. Neuen BL, Bello AK, Levin A, Lunney M, Osman MA, Ye F, et al. National health policies and strategies for addressing chronic kidney disease: Data from the International Society of Nephrology Global Kidney Health Atlas. PLOS Glob Public Health. 2023;3(2):e0001467.

30. Aregawi K, Kabew Mekonnen G, Belete R, Kucha W. Prevalence of chronic kidney disease and associated factors among adult diabetic patients: a hospital-based cross-sectional study. Frontiers in epidemiology. 2024;4:1467911.

31. Israel E, Borko UD, Mota K, Tesfaw M, Feleke T, Abraham A, et al. Out of sight: chronic kidney diseases among diabetic patients attending care and follow up. Findings from pastoralist health facilities of Southern Ethiopia. Front Public Health. 2024;12:1326011.

32. Damtie S, Biadgo B, Baynes HW, Ambachew S, Melak T, Asmelash D, et al. Chronic Kidney Disease and Associated Risk Factors Assessment among Diabetes Mellitus Patients at A Tertiary Hospital, Northwest Ethiopia. Ethiop J Health Sci. 2018;28(6):691–700.

33. Abdulkadr M, Merga H, Mizana BA, Terefe G, Dube L. Chronic Kidney Disease and Associated Factors among Diabetic Patients at the Diabetic Clinic in a Police Hospital, Addis Ababa. Ethiop J Health Sci. 2022;32(2):307–12.

34. Bahrey D, Gebremedhn G, Mariye T, Girmay A, Aberhe W, Hika A, et al. Prevalence and associated factors of chronic kidney disease among adult hypertensive patients in Tigray teaching hospitals: a cross-sectional study. BMC Res Notes. 2019;12(1):562.

35. Abuhay HW, Yenit MK, Melese M, Alemu GG, Aragaw FM. Prevalence and associated factors of chronic kidney disease among diabetes mellitus patients in Ethiopia: A systematic review and meta-analysis. PLoS One. 2025;20(3):e0315529.

36. Cheru A, Edessa D, Regassa LD, Gobena T. Incidence and predictors of chronic kidney disease among patients with diabetes treated at governmental hospitals of Harari Region, eastern Ethiopia, 2022. Front Public Health. 2023;11:1290554.

37. Afkarian M, Zelnick LR, Hall YN, Heagerty PJ, Tuttle K, Weiss NS, et al. Clinical Manifestations of Kidney Disease Among US Adults With Diabetes, 1988-2014. Jama. 2016;316(6):602–10.

38. Francis A, Harhay MN, Ong ACM, Tummalapalli SL, Ortiz A, Fogo AB, et al. Chronic kidney disease and the global public health agenda: an international consensus. Nature reviews Nephrology. 2024;20(7):473–85.

39. Hirst JA, Hill N, O’Callaghan CA, Lasserson D, McManus RJ, Ogburn E, et al. Prevalence of chronic kidney disease in the community using data from OxRen: a UK population-based cohort study. The British journal of general practice : the journal of the Royal College of General Practitioners. 2020;70(693):e285–e93.

40. Bello AK, McIsaac M, Okpechi IG, Johnson DW, Jha V, Harris DCH, et al. International Society of Nephrology Global Kidney Health Atlas: structures, organization, and services for the management of kidney failure in North America and the Caribbean. Kidney international supplements. 2021;11(2):e66–e76.

41. Herrera-Añazco P, Taype-Rondan A, Lazo-Porras M, Alberto Quintanilla E, Ortiz-Soriano VM, Hernandez AV. Prevalence of chronic kidney disease in Peruvian primary care setting. BMC Nephrol. 2017;18(1):246.

42. Liyanage T, Toyama T, Hockham C, Ninomiya T, Perkovic V, Woodward M, et al. Prevalence of chronic kidney disease in Asia: a systematic review and analysis. BMJ Glob Health. 2022;7(1).

43. Hill NR, Fatoba ST, Oke JL, Hirst JA, O’Callaghan CA, Lasserson DS, et al. Global Prevalence of Chronic Kidney Disease - A Systematic Review and Meta-Analysis. PLoS One. 2016;11(7):e0158765.

44. George C, Stoker S, Okpechi I, Woodward M, Kengne A. The Chronic Kidney Disease in Africa (CKD-Africa) collaboration: lessons from a new pan-African network. BMJ Glob Health. 2021;6(8).

45. Wagnew F, Eshetie S, Kibret GD, Zegeye A, Dessie G, Mulugeta H, et al. Diabetic nephropathy and hypertension in diabetes patients of sub-Saharan countries: a systematic review and meta-analysis. BMC Res Notes. 2018;11(1):565.

46. Kibirige D, Sekitoleko I, Lumu W. Burden and predictors of diabetic kidney disease in an adult Ugandan population with new-onset diabetes. BMC Res Notes. 2023;16(1):234.

47. Shiferaw WS, Akalu TY, Aynalem YA. Chronic Kidney Disease among Diabetes Patients in Ethiopia: A Systematic Review and Meta-Analysis. International journal of nephrology. 2020;2020:8890331.

48. Fiseha T, Ahmed E, Chalie S, Gebreweld A. Prevalence and associated factors of impaired renal function and albuminuria among adult patients admitted to a hospital in Northeast Ethiopia. PLoS One. 2021;16(2):e0246509.

49. Tekalign T, Guta MT, Awoke N, Chichiabellu TY, Meskele M, Anteneh G, et al. Time to Diabetic Nephropathy and its Predictors Among Diabetic Patients Treated in Wolaita and Dawuro Zone Hospitals, Ethiopia: A Retrospective Cohort Study. International journal of nephrology and renovascular disease. 2023;16:163-72.

50. Neale EP, Middleton J, Lambert K. Barriers and enablers to detection and management of chronic kidney disease in primary healthcare: a systematic review. BMC Nephrol. 2020;21(1):83.

51. Seleshi T, Alemneh T, Mekonnen D, Tesfaye D, Markos S, Getachew Y, et al. Assessment of subclinical left ventricular systolic and diastolic dysfunction in patients with type 2 diabetes mellitus under follow-up at Tikur Anbessa specialized hospital, Ethiopia: a case-control study. BMC cardiovascular disorders. 2024;24(1):201.

52. Alemu H, Hailu W, Adane A. Prevalence of Chronic Kidney Disease and Associated Factors among Patients with Diabetes in Northwest Ethiopia: A Hospital-Based Cross-Sectional Study. Current therapeutic research, clinical and experimental. 2020;92:100578.

53. Matsha TE, Erasmus RT. Chronic kidney disease in sub-Saharan Africa. The Lancet Global health. 2019;7(12):e1587–e8.

54. Fan Z, Yun J, Yu S, Yang Q, Song L. Alcohol Consumption Can be a "Double-Edged Sword" for Chronic Kidney Disease Patients. Medical science monitor : international medical journal of experimental and clinical research. 2019;25:7059–72.

55. Li Y, Zhu B, Song N, Shi Y, Fang Y, Ding X. Alcohol consumption and its association with chronic kidney disease: Evidence from a 12-year China health and Nutrition Survey. Nutrition, metabolism, and cardiovascular diseases : NMCD. 2022;32(6):1392–401.

56. Jha V, Garcia-Garcia G, Iseki K, Li Z, Naicker S, Plattner B, et al. Chronic kidney disease: global dimension and perspectives. Lancet. 2013;382(9888):260-72.

57. Mwenda V, Githuku J, Gathecha G, Wambugu BM, Roka ZG, Ong’or WO. Prevalence and factors associated with chronic kidney disease among medical inpatients at the Kenyatta National Hospital, Kenya, 2018: a cross-sectional study. Pan Afr Med J. 2019;33:321.

58. Machado-Duque ME, Gaviria-Mendoza A, Valladales-Restrepo LF, Franco JS, de Rosario Forero M, Vizcaya D, et al. Treatment patterns of antidiabetic and kidney protective therapies among patients with type 2 diabetes mellitus and chronic kidney disease in Colombia. The KDICO descriptive study. Diabetol Metab Syndr. 2023;15(1):150.

